# Health Security Across the Spectrum: Exploring the Impact of Socioeconomic Factors on Health Insurance Uptake in India

**DOI:** 10.1101/2024.06.19.24309161

**Authors:** Madhubrota Chatterjee, Alok Aditya, Prashant Kumar Choudhary

**Affiliations:** Population Research Centre, Institute for Social and Economic Change, Bengaluru, India; Centre for Economic Studies and Policy, Institute for Social and Economic Change, Bengaluru, India; Department of Public Policy, Manipal Academy of Higher Education, Bengaluru, India

**Keywords:** Health, Health insurance choice, Diseases, Older people, Services, Access

## Abstract

To ensure that healthcare services are accessible, health insurance is receiving increased amounts of attention in debates among health experts. The current disease pattern has forced people to rely on intensive care, which has increased both the cost of treatment and the frequency of accessing healthcare facilities. This paper has focused on various socioeconomic and demographic determinants of choice that are important for ensuring the use of various types of health insurance schemes or programs among the older population in India. Using the Longitudinal Survey of Aging in India (LASI) Wave-1 2020 data, the total sample used for this study included 66,658 elderly individuals aged 45 years and older. Both binary association and multinomial logistic regression were employed to examine the associations of all the socioeconomic and demographic determinants with people having and not having access to different health insurance. The findings showed that there is a greater incidence of government health insurance than of community, employer, or private health insurance among people in all categories based on demographic, geographical, economic distribution, and health status, with chronic diseases showing some impact on people choosing to be insured. This study recommends that policy actions be taken to make the health insurance market transparent and reduce the chances of failure.

**Contributions to the literature:**

- The current epidemiological transition, along with the prospective demographic transition of more people in later stages of life that India is currently facing and will experience in the coming decades, has raised severe concerns about the accessibility of healthcare services.
- Knowing the factors and reasons for choosing certain health insurance schemes is highly important for tracing the actual lags in the Indian health insurance market and will help policymakers formulate health insurance policies.
- The need to make the Indian health insurance market transparent.
- reducing the chances of failure of health insurance schemes

## BACKGROUND

The accessibility of healthcare to a certain extent is dependent on health insurance coverage. Health insurance has been gaining attention in recent debates among health experts and policymakers. The general rise in the price of medical tools and medicines and the rise in the cost of care have created serious issues in the accessibility of healthcare services. The current epidemiological pattern of various diseases has forced people to rely on multispecialty hospitals and intensive care medical facilities. Consequently, both the cost of treatment and the frequency of visiting healthcare facilities have increased. In countries such as India, health reform is considered a major government agenda, and health insurance promotes health reform and the government’s poverty reduction agenda (Reddy, 2012). However, the Indian public healthcare system is often characterized by mismanagement, pro-rich status (Berman, 2007), poor access to and quality of service, and high response time. In such cases, people are now moving toward private healthcare facilities. According to the NSSO (various rounds) data, in 2017-2018, the private sector provided 54 and 65 percent of hospitalization care in rural and urban India, respectively. Moreover, 67 percent and 74 percent of outpatient care is provided by the private sector in both rural and urban India, respectively. Hence, India is one of the countries with the highest levels of out-of-pocket expenditure (OPE) in the world, where 60% of the total healthcare expenditure consists of out-of-pocket expenditure. The impact can be reduced to 30 percent if the government increases public healthcare expenditures to 3 percent of GDP. However, currently, India ranks 179^th^ among 189 countries in terms of prioritizing health in government budgets.

According to the 2011 census of India, 104 million people aged >60 years constitute 8.6% of the total population, and this number is projected to increase by 157 million in 2025 and 297 million in 2050, which will constitute 11 and 18%, respectively, of the total population (Giridhar *et al*., 2014). The change in the demographic structure in India has significant socioeconomic and public health implications, and the current epidemiological transition in India leads to these implications. Non-communicable diseases are now new emerging characteristics of the health burden of the Indian population (Cramm *et al*., 2015). An increasing share of the older population leads to increased demand for health services, but vast inequality in accessing economic and social resources leads to inadequate access to healthcare services (Lopreite & Mauro, 2017). The social and economic gap due to modernization and globalization has intensified competition among the poor section of the older population accessing socioeconomic resources (Bhat & Dhruvarajan, 2002). An increase in health problems among older people leads to large expenditures on health, which in turn negatively affects the consumption of other goods and services (Pal, 2013; Joglekar, 2012) and increases the chance of being pushed into indebtedness, leading to poverty (Joglekar, 2012; Damme *et al*., 2004). Hence, the increasing demand and cost of healthcare services, unequal accessibility, and resource constraints raise concerns about alternative sources of financing healthcare expenditures. Although public subsidies have reduced out-of-pocket expenditures, they favor maternity and child healthcare only (Berman, 2007). Therefore, hedging health risk through health insurance or other prepaid schemes has become an important tool for health reform because it not only improves the provisions for healthcare services but also improves the quality and efficiency of the healthcare system (Ahuja, 2004). However, two problems facing the provision of comprehensive and inclusive health insurance for Indian health experts are first, the way to convert private out-of-pocket expenditures into insurance premiums from a large number of insured people, and second, how to access health insurance for people who are not able to pay the full amount of the premium. In the late 1990s, the government started community-based health insurance schemes that sought to provide better and more access to health insurance at the village level. Community-based health insurance is more suitable for providing health insurance to the poor population because it can take different forms based on the characteristics of the targeted population (Ahuja, 2004). However, community-based health insurance schemes are not free from criticism, as these schemes only partially cover some specific health risks. Therefore, to make it attractive to clients, insurance must cover their prospective needs and should also be affordable (Radwan, 2005; Danis *et al*., 2007).

A negative health shock on an individual incurs two types of costs, i.e., the direct cost associated with impoverished health and reduced productivity, which also affects the individual earnings and out-of-pocket expenditures associated with the treatment of the disease (Liu, 2015). To cope with this negative health shock, individuals without an insurance plan use different channels of self-insurance to smooth household income, while access to health insurance lessens the impact of negative health shock by hedging individual earnings and providing access to high-quality health services (Liu, 2015); additionally, they better utilize these services (Arokiasamy & Srinivas, 2013; Prinja, 2019; Awoke *et al*., 2017; Thuong *et al*, 2020); and lower out-of-pocket health expenditures (Thuong *et al*, 2020; Fan *et al*., 2012; Dror, 2006; Kumar, 2011).

The Indian health insurance market is facing the problems of both asymmetric information and low efficiency of service delivery. As Akerlof (1970) noted, asymmetric information leads to uncertainty about the quality of the product, and consequently, only a low-quality product is delivered to the market. The fear of adverse selection among insurance companies leads to the problems of inefficient delivery, low coverage, and high premiums; consequently, the market remains underdeveloped. The public health insurance programs in India can be categorized into three distinct generations based on distinct governance modes. First, privately funded public service delivery leads to underutilization and poor service quality; second, publicly funded service delivery contracts to the private sector also result in underperformance and low quality due to the malfunctioning private sector; and the third generation continues the publicly funded program, but in service delivery, it prefers public-sector agencies that are yet to be assessed (Maurya, 2019). The entry of private sector insurance companies after liberalization did not cause any significant positive changes in the volume, products, or delivery systems of health insurance (Ahuja, 2004; Mavalankar & Bhat, 2000). Thus, supply-side inefficiency and noncompetitiveness result in low demand for insurance. The myth that poor people do not show interest in purchasing insurance benefits is challenged by the findings of Dror (2006), who, based on household survey data collected in seven locations with micro health insurance units under operation, show that most people are willing to pay 1 percent or more of their total income as a premium for their health insurance, while the poorest section of society prioritizes access to some form of healthcare. They are willing to pay a higher percentage of their household income. Most households prefer comprehensive benefit health insurance packages rather than partial insurance packages (Mathiyazhagan, 1998; Dror, 2007). People prefer high-probability- and low-cost care over low-probability- and high-cost care when they choose health insurance (Dror *et al*., 2007). Dror *et al*. (2007) suggested that to encourage rural, poor, and illiterate households with very little knowledge of health insurance, benefit packages should be offered with a limited budget of Rs.500 per household per year. The World Bank (2001) considers low-frequency and high-unit-cost events to be the cause of deterioration of health and health-related services. However, this view was challenged by Danis *et al*. (2007), who found that benefit packages that provide better access to health services (neither catastrophic nor rare) and can accumulate to prohibitive financial burdens are chosen by participants.

The choice of being insured with health insurance depends on the socioeconomic and demographic characteristics of the participants. Studies show that among the various socioeconomic determinants of health insurance occupation, income, education, health expenditure, and awareness (Sukumar, 2009; Isabella, 2011; Ramarshna, 2012; Arokiasamy & Srinivas, 2013; Ahire & Rishipathak, 2019), employment status and household size (Isabella, 2011), and cast and religious affiliation (Arokiasamy & Srinivas, 2013) have significant impacts on the choice of being insured with health insurance. The availability of healthcare facilities, trust in insurance companies (Ahire & Rishipathak, 2019), rural ◻ urban disparities (Arokiasamy & Srinivas, 2013), and gender disparities in the utilization of health insurance, mostly in children and elderly individuals (Dupas & Jain, 2021), have also been found to affect the choice to be insured with health insurance. Dror *et al*. (2006) found that willingness to pay (WTP) for insurance and household size are positively and proportionally correlated, while although the WTP and household income are positively correlated, they decrease with increasing income. They also found a positive correlation between WTP and the educational level of the head of the household.

The choice of health insurance becomes a very important issue when we talk about the older section of the population. Availing health insurance for the older population significantly improves health status (Baker *et al*., 2006) beyond age 65 and possibly reduces the medical spending of each new group of 65-year-old medicare beneficiaries (Hadley & Waidmann, 2006). This approach not only increases the number of insured people but also creates potential savings for public insurance programs (the study was based on providing insurance to Americans aged between 55 and 64 years). The studies show mixed results concerning the impact of age or increasing age on the choice and WTP for health insurance. However, the results differ from place to place. Several studies have shown that age and increasing age do not affect the choice of health insurance (Ramarshna, 2012; case study based on Hyderabad city in India) and do not affect the WTP for health insurance (Dror *et al*., 2006); however, other studies have shown that age and increasing age significantly impact both the WTP for insurance (Becker & Zweifel, 2008) and the choice to be insured (Bhat & Jain, 2006; Becker & Zweifel, 2008).

This paper will focus on the possible socioeconomic, demographic, and health factors affecting the choice of the older population to be insured with various health insurance. We considered the choice of four categories of health insurance schemes or programs, i.e., government, employer, community, and private. This study is based on longitudinal aging survey data from the older population aged 45 years and older.

## METHODOLOGY

### Data source

We used data from the available Longitudinal Aging Study of India-Wave 1. The LASSO study is the world’s largest and largest longitudinal aging study in India; it was funded by the Ministry of Health and Family Welfare, the Government of India, the National Institute on Aging, and the United Nations Population Fund, India. This was a collaborative study of three main partnering institutions: the International Institute for Population Sciences (IIPS), Harvard T.H. Chan School of Public Health (HSPH), the University of Southern California (USC), and several other national and international institutions. This was a panel study of 72,250 older adults aged 45 and above, including spouses aged less than 45 years, representatives of India, and all of its states and union territories. The LASSO is designed to provide researchers and policymakers with comprehensive data on the key economic, social, and health characteristics of India’s older population. The survey instrument comprises the household survey schedule; individual survey schedule; and biomarker surveys, including questions about demographic, economic, employment, behavioral, social, physical, and mental health characteristics; and an extensive set of cognitive functioning tests. The survey design, tools, and protocols are consistent with those of the Health and Retirement Study (HRS) in the U.S. and its sister surveys in Asia, Europe, and elsewhere (IIPS, 2020).

### Variables

#### Outcome Variable

This study included 66,658 adults aged 45 years and older. Participants in the survey were asked to mention the types of health insurance they access. The responses were recoded into five categories: not having any health insurance, providing government insurance, providing community insurance, providing employer insurance, and providing private insurance.

#### Explanatory Variables

##### Sociodemographic Factors

The variables included were sex, age group, place of residence, region, religion, caste, marital status, living arrangement, and education. Marital status was coded as currently married/living together or single (including widowed, separated, deserted and divorced individuals).

##### Socioeconomic

The socioeconomic measures included were household income/wealth quintile (coded as 0 for rich, 1 for middle, and 2 for poor), working status coded as never worked, currently working, currently not working, and retired. An index of having access to Basic Amenities was created that covers households with cooking fuel, toilets, water, and electricity.

##### Health

In this category, self-reported health status (coded as Very Good, Good, Fair, Poor, or Very Poor) and chronic morbidity (with any one of the following conditions: hypertension, diabetes, cancer, lung disease, heart disease, stroke, arthritis, bone disease, neurological disease, or cholesterol or not) were included. The physical activity status was also considered.

Additionally, variables such as natural and man-made disasters were taken to determine the effects of these disasters on claims of insurance.

All the categorical explanatory variables are coded according to the LASI framework.

### Statistical analysis

To examine the association between the four types of health insurance and the responsible determinants, cross-tabulation was performed by organizing them in a table in percentage form, which is known as bivariate analysis. Bivariate analysis is the simultaneous analysis of two variables (attributes). It is used to explore the concept of a relationship between two variables, whether there exists an association and the strength of this association, or whether there are differences between two variables and the significance of these differences.

Since the outcome variable in our study, Health Insurance, has more than two categories, multinomial logistic regression was used. It is used to model nominal outcome variables in which the log odds of the outcomes are modeled as a linear combination of the predictor variables. In this paper, this metric is shown in the form of the relative risk ratio. The ratio of the probability of choosing one outcome category to the probability of choosing the baseline category is often referred to as the relative risk. Like the odds ratio, relative risk can be obtained by exponentiating the linear equations above, yielding regression coefficients that are relative risk ratios for a unit change in the predictor variable (Introduction to SAS: UCLA, 2021).

Statistical analyses were conducted using Stata 16, version SE.

## RESULTS

Table 1 presents the descriptive statistics of the study sample. Throughout the study sample of this paper, people have opted for having no health insurance, but if this is the case, government insurance tops the list among the people providing it across several possible determinants. Across age groups, government insurance is more common among people younger than 60 years of age, i.e., 21 percent, followed by people in the 60-69 age group, which is 20 percent. Most females have no health insurance, but their choice of having community health insurance is greater (0.21 percent) than that of males (0.19 percent). People residing in urban areas have more employer health insurance (1.98 percent) and private health insurance (2.62 percent), whereas people in urban areas have a greater prevalence of government health insurance (22.27 percent). Across regions, government insurance is more prevalent in the northeast (28.76%), community insurance is more prevalent in eastern India (0.79%), employer insurance is more prevalent in northern regions (1.46%), and private insurance is more prevalent in western India (2.43%) than in their respective counterparts. A higher percentage of Christians (27.92%) and people from the Scheduled Tribe Community (29.03%) had government insurance than did the remaining people. People currently married and living with spouses had more government insurance, which was available for 20 percent and 20.18 percent, respectively. Those with a college degree and above and who are classified as rich in household wealth are more inclined to use private insurance, with shares of 7.54 percent and 2.26 percent, respectively, than are the remaining two groups. Individuals with very good self-rated health, without any morbidity, and who never engaged in physical activities tend to have no health insurance.

**Table 1:** Description of the study sample (sociodemographic, socioeconomic, and health characteristics of adults) by their choice of type of health insurance.

|  | No Insurance | Government | Community | Employer | Private | Total |
| --- | --- | --- | --- | --- | --- | --- |
| **Age** |  |  |  |  |  |  |
| Adult (45-59) | 28322 (76.09) | 7664 (20.59) | 88 (0.24) | 530 (1.42) | 619 (1.66) | **37223 (100)** |
| Young-old (60-69) | 14036 (77.65) | 3584 (19.83) | 33 (0.18) | 185 (1.02) | 238 (1.32) | **18076 (100)** |
| Older-Old (70-79) | 5510 (80.16) | 1228 (17.86) | 4 (0.06) | 53 (0.77) | 79 (1.15) | **6874 (100)** |
| Oldest-old (80 & above) | 3977 (88.67) | 463 (10.32) | 4 (0.09) | 24 (0.54) | 17 (0.38) | **4485 (100)** |
| **Sex** |  |  |  |  |  |  |
| Male | 23219 (76.18) | 6197 (20.33) | 57 (0.19) | 493 (1.62) | 513 (1.68) | **30479 (100)** |
| Female | 27530 (78.47) | 6742 (19.22) | 72 (0.21) | 299 (0.85) | 440 (1.25) | **35083 (100)** |
| **Residence** |  |  |  |  |  |  |
| Rural | 32681 (75.98) | 9580 (22.27) | 95 (0.22) | 324 (0.75) | 334 (0.78) | **43014 (100)** |
| Urban | 19164 (81.05) | 3359 (14.21) | 34 (0.14) | 468 (1.98) | 619 (2.62) | **23644 (100)** |
| **Region** |  |  |  |  |  |  |
| North | 14763 (89.16) | 1380 (8.33) | 11 (0.07) | 242 (1.46) | 162 (0.98) | **16558 (100)** |
| South | 12208 (70.96) | 4319 (25.1) | 20 (0.12) | 246 (1.43) | 411 (2.39) | **17204 (100)** |
| East | 8916 (76.1) | 2512 (21.44) | 93 (0.79) | 110 (0.94) | 85 (0.73) | **11716 (100)** |
| Central | 3684 (78.57) | 852 (18.17) | 4 (0.09) | 61 (1.3) | 88 (1.88) | **4689 (100)** |
| West | 6172 (78.59) | 1392 (17.73) | 1 (0.01) | 97 (1.24) | 191 (2.43) | **7853 (100)** |
| North East | 6102 (70.64) | 2484 (28.76) | 0 (0) | 36 (0.42) | 16 (0.19) | **8638 (100)** |
| **Religion** |  |  |  |  |  |  |
| Hindu | 36671 (76.24) | 9861 (20.5) | 99 (0.21) | 684 (1.42) | 784 (1.63) | **48099 (100)** |
| Muslim | 6672 (85.51) | 1002 (12.84) | 25 (0.32) | 46 (0.59) | 58 (0.74) | **7803 (100)** |
| Christian | 4598 (70.35) | 1825 (27.92) | 1 (0.02) | 42 (0.64) | 70 (1.07) | **6536 (100)** |
| Others | 2701 (90.39) | 226 (7.56) | 3 (0.1) | 20 (0.67) | 38 (1.27) | **2988 (100)** |
| **Caste Category** |  |  |  |  |  |  |
| General | 13572 (83.37) | 1957 (12.02) | 32 (0.2) | 294 (1.81) | 424 (2.6) | **16279 (100)** |
| Scheduled caste | 8722 (79.59) | 2006 (18.3) | 35 (0.32) | 131 (1.2) | 65 (0.59) | **10959 (100)** |
| Scheduled tribe | 7847 (69.05) | 3299 (29.03) | 15 (0.13) | 74 (0.65) | 130 (1.14) | **11365 (100)** |
| Other backward class (OBC) | 18771 (76.22) | 5242 (21.28) | 34 (0.14) | 276 (1.12) | 306 (1.24) | **24629 (100)** |
| **Marital Status** |  |  |  |  |  |  |
| Currently Married | 37436 (76.76) | 9756 (20) | 96 (0.2) | 686 (1.41) | 795 (1.63) | **48769 (100)** |
| Widowed/Divorced/Separated | 12347 (79.29) | 2957 (18.99) | 31 (0.2) | 95 (0.61) | 141 (0.91) | **15571 (100)** |
| Others | 964 (79.08) | 225 (18.46) | 2 (0.16) | 11 (0.9) | 17 (1.39) | **1219 (100)** |
| **Living Arrangement** |  |  |  |  |  |  |
| Alone | 1839 (79.51) | 433 (18.72) | 5 (0.22) | 12 (0.52) | 24 (1.04) | **2313 (100)** |
| With Spouse | 36658 (76.57) | 9663 (20.18) | 96 (0.2) | 677 (1.41) | 783 (1.64) | **47877 (100)** |
| With Children | 9816 (78.9) | 2398 (19.27) | 22 (0.18) | 87 (0.7) | 118 (0.95) | **12441 (100)** |
| Others | 2436 (83.11) | 445 (15.18) | 6 (0.2) | 16 (0.55) | 28 (0.96) | **2931 (100)** |
| **Education** |  |  |  |  |  |  |
| Less than Primary | 5189 (69.39) | 2150 (28.75) | 19 (0.25) | 56 (0.75) | 64 (0.86) | **7478 (100)** |
| Primary | 11412 (76.8) | 3024 (20.35) | 21 (0.14) | 209 (1.41) | 193 (1.3) | **14859 (100)** |
| Secondary | 4653 (79.07) | 961 (16.33) | 9 (0.15) | 122 (2.07) | 140 (2.38) | **5885 (100)** |
| Higher Secondary | 2453 (78.47) | 456 (14.59) | 11 (0.35) | 88 (2.82) | 118 (3.77) | **3126 (100)** |
| College and above | 2559 (75.42) | 408 (12.02) | 5 (0.15) | 165 (4.86) | 256 (7.54) | **3393 (100)** |
| **Household Wealth Quintile** |  |  |  |  |  |  |
| Rich | 20844 (78.23) | 4675 (17.55) | 55 (0.21) | 468 (1.76) | 603 (2.26) | **26645 (100)** |
| Middle | 10422 (77.78) | 2684 (20.03) | 26 (0.19) | 140 (1.04) | 128 (0.96) | **13400 (100)** |
| Poor | 20579 (77.33) | 5580 (20.97) | 48 (0.18) | 184 (0.69) | 222 (0.83) | **26613 (100)** |
| **Basic Amenities Index** |  |  |  |  |  |  |
| 0 | 1018 (76.66) | 275 (20.71) | 1 (0.08) | 16 (1.2) | 18 (1.36) | **1328 (100)** |
| 0.25 | 2132 (84.77) | 365 (14.51) | 2 (0.08) | 6 (0.24) | 10 (0.4) | **2515 (100)** |
| 0.5 | 7566 (76.91) | 2140 (21.75) | 19 (0.19) | 50 (0.51) | 63 (0.64) | **9838 (100)** |
| 0.75 | 17238 (74.21) | 5517 (23.75) | 61 (0.26) | 199 (0.86) | 213 (0.92) | **23228 (100)** |
| 1 | 23887 (80.31) | 4642 (15.61) | 46 (0.15) | 521 (1.75) | 649 (2.18) | **29745 (100)** |
| **Working Status** |  |  |  |  |  |  |
| Never Worked | 14956 (83.15) | 2640 (14.68) | 34 (0.19) | 132 (0.73) | 224 (1.25) | **17986 (100)** |
| Currently Working | 22261 (73.77) | 6908 (22.89) | 66 (0.22) | 452 (1.5) | 490 (1.62) | **30177 (100)** |
| Currently Not Working | 11507 (77.7) | 3007 (20.3) | 24 (0.16) | 103 (0.7) | 169 (1.14) | **14810 (100)** |
| Retired | 2011 (78.16) | 382 (14.85) | 5 (0.19) | 105 (4.08) | 70 (2.72) | **2573 (100)** |
| **Self-Rated Health** |  |  |  |  |  |  |
| Very Good | 2461 (80.69) | 428 (14.03) | 6 (0.2) | 65 (2.13) | 90 (2.95) | **3050 (100)** |
| Good | 18094 (76.58) | 4814 (20.37) | 25 (0.11) | 335 (1.42) | 360 (1.52) | **23628 (100)** |
| Fair | 21060 (78.22) | 5172 (19.21) | 58 (0.22) | 274 (1.02) | 360 (1.34) | **26924 (100)** |
| Poor | 7437 (75.8) | 2113 (21.54) | 34 (0.35) | 103 (1.05) | 124 (1.26) | **9811 (100)** |
| Very Poor | 919 (73.99) | 290 (23.35) | 6 (0.48) | 10 (0.81) | 17 (1.37) | **1242 (100)** |
| **Morbid Condition** |  |  |  |  |  |  |
| No Morbidity | 28228 (78.19) | 7054 (19.54) | 60 (0.17) | 352 (0.98) | 408 (1.13) | **36102 (100)** |
| Morbidity | 13849 (76.94) | 3584 (19.91) | 42 (0.23) | 255 (1.42) | 270 (1.5) | **18000 (100)** |
| Multimorbidity | 9753 (77.82) | 2292 (18.29) | 27 (0.22) | 185 (1.48) | 275 (2.19) | **12532 (100)** |
| **Physical Activities Status** |  |  |  |  |  |  |
| Never | 30923 (78.68) | 7255 (18.46) | 70 (0.18) | 455 (1.16) | 599 (1.52) | **39302 (100)** |
| Often | 7539 (74.61) | 2303 (22.79) | 22 (0.22) | 132 (1.31) | 109 (1.08) | **10105 (100)** |
| Frequently | 11720 (75.2) | 3379 (21.68) | 37 (0.24) | 205 (1.32) | 245 (1.57) | **15586 (100)** |
| **Natural Disaster** |  |  |  |  |  |  |
| No | 49323 (77.26) | 12662 (19.83) | 127 (0.2) | 786 (1.23) | 940 (1.47) | **63838 (100)** |
| Yes | 1225 (80.43) | 277 (18.19) | 2 (0.13) | 6 (0.39) | 13 (0.85) | **1523 (100)** |

| **Man-made Disaster** |  |  |  |  |  |  |
| --- | --- | --- | --- | --- | --- | --- |
| No | 49947 (77.34) | 12792 (19.8) | 128 (0.2) | 778 (1.2) | 939 (1.45) | **64584 (100)** |
| Yes | 599 (77.29) | 147 (18.97) | 1 (0.13) | 14 (1.81) | 14 (1.81) | **775 (100)** |
| **Total** | **51845 (77.78)** | **12939 (19.41)** | **129 (0.19)** | **792 (1.19)** | **953 (1.43)** | **66658 (100)** |

Table 2 shows the results from the multinomial logistic regression in the form of relative risk ratios. Across all age groups, the oldest old individuals had a significantly lower risk of receiving government, employer, and private health insurance than adults did (RRRs= 0.572, 0.552, and 0.331, respectively). Urban people have a significantly greater risk of having both employer (RRR= 1.65) and private health insurance (RRR= 1.993) and lower government health insurance (RRR=0.665) than do people in rural areas. Across regions, individuals residing in both Eastern and Southern India have greater risks of receiving community health insurance (RRRs= 13.842 and 4.053, respectively) than do those residing in North India. As reflected in the first table itself, people from Western regions are 2.5 times more likely to have private health insurance, and people from Northeast regions are 3.2 times more likely to have government health insurance. Among caste categories, scheduled castes, scheduled tribes, and other backward casts have shown higher risks of having government health insurance (RRRs= 1.12, 1.43, and 1.27, respectively) than does the general (unreserved) category. Those with a college degree and above are 2.9 times more likely to have employer health insurance and 4 times more likely to have private health insurance than are those with no education. Similarly, people with access to basic amenities such as toilets, drinking water, electricity, and cooking fuel are 1.5 times more likely to have access to private health insurance. Those who were still working and those who retired had greater odds of having employer health insurance (RRR= 2.18 and 3.64, respectively) than did those who had never worked. People are more likely to have employer health insurance, followed by government insurance. Those who reported their health status to be “very poor” were 2 times more likely to use government health insurance, and individuals with multimorbidity were at greater risk for having private health insurance (RRR= 1.477) than were those with zero morbidity. Individuals affected by natural disasters are 0.26 times less likely to have employer health insurance; conversely, those impacted by man-made disasters are 1.91 times more likely to be covered by employer health insurance.

**Table 2:** Results of the multinomial logistic regression analysis of the types of health insurance across all the sociodemographic, socioeconomic, and health determinants.

|  | **Government** |  | **Community** |  | **Employer** |  | **Private** |  |
| --- | --- | --- | --- | --- | --- | --- | --- | --- |
| **Determinants** | **RRR** | **St. Err.** | **RRR** | **St. Err.** | **RRR** | **St. Err.** | **RRR** | **St. Err.** |
| **Age: Adult<sup>®</sup>** |  |  |  |  |  |  |  |  |
| Young-old | 0.907*** | 0.033 | 0.423** | 0.157 | 0.713*** | 0.082 | 0.877 | 0.089 |
| Older-Old | 0.785*** | 0.045 | 0.075** | 0.078 | 0.561*** | 0.107 | 0.584*** | 0.102 |
| Oldest-old | 0.572*** | 0.054 | 0 | 0 | 0.552* | 0.176 | 0.331*** | 0.124 |
| **Sex: Male<sup>®</sup>** |  |  |  |  |  |  |  |  |
| Female | 1.041 | 0.041 | 0.673 | 0.278 | 0.989 | 0.117 | 0.986 | 0.106 |
| **Residence: Rural®** |  |  |  |  |  |  |  |  |
| Urban | 0.665*** | 0.022 | 0.627 | 0.197 | 1.65*** | 0.16 | 1.993*** | 0.189 |
| **Region: Northern®** |  |  |  |  |  |  |  |  |
| South | 3.327*** | 0.171 | 4.053** | 2.246 | 1.196 | 0.138 | 3.219*** | 0.377 |
| East | 2.704*** | 0.146 | 13.842*** | 6.982 | 0.81 | 0.112 | 1.106 | 0.168 |
| Central | 1.546*** | 0.119 | 1.932 | 1.637 | 1.153 | 0.205 | 2.091*** | 0.365 |
| West | 1.787*** | 0.111 | 0.405 | 0.443 | 0.961 | 0.132 | 2.549*** | 0.326 |
| North East | 3.204*** | 0.197 | 0 | 0.001 | 0.271*** | 0.068 | 0.242*** | 0.073 |
| **Religion: Hindu®** |  |  |  |  |  |  |  |  |
| Muslim | 0.485*** | 0.029 | 1.437 | 0.571 | 0.45*** | 0.083 | 0.352*** | 0.063 |
| Christian | 1.085 | 0.06 | 0.367 | 0.381 | 0.688* | 0.148 | 0.867 | 0.145 |
| Others | 0.453*** | 0.047 | 3.027 | 2.055 | 0.454*** | 0.11 | 1.101 | 0.202 |
| **Caste: General®** |  |  |  |  |  |  |  |  |
| Scheduled caste | 1.121** | 0.057 | 1.388 | 0.527 | 1.14 | 0.151 | 0.41*** | 0.069 |
| Scheduled tribe | 1.431*** | 0.079 | 1.377 | 0.712 | 1.306 | 0.231 | 1.095 | 0.178 |
| Other backward class | 1.27*** | 0.049 | 0.69 | 0.24 | 0.892 | 0.092 | 0.579*** | 0.054 |
| **Marital Status: Currently Married®** |  |  |  |  |  |  |  |  |
| Widowed/Divorced/Separated | 1.898*** | 0.308 | 2094934.7 | 2.5E+09 | 1.165 | 0.488 | 1.211 | 0.45 |
| Others | 1.645*** | 0.316 | 0.822 | 1256.504 | 1.453 | 0.747 | 1.267 | 0.573 |
| **Living arrangement: Living alone®** |  |  |  |  |  |  |  |  |
| With Spouse | 2.33*** | 0.412 | 952084.73 | 1.14E+09 | 1.728 | 0.769 | 2.238* | 0.945 |
| With Children | 1.188* | 0.123 | 0.279 | 0.259 | 1.124 | 0.376 | 1.441 | 0.471 |
| Others | 0.904 | 0.119 | 1.098 | 1.121 | 0.714 | 0.308 | 1.08 | 0.425 |
| **Education: No Education®** |  |  |  |  |  |  |  |  |
| Primary | 0.729*** | 0.026 | 0.518* | 0.183 | 1.379** | 0.215 | 1.098 | 0.165 |
| Secondary | 0.613*** | 0.03 | 0.549 | 0.251 | 1.564*** | 0.267 | 1.457** | 0.234 |
| Higher Secondary | 0.559*** | 0.035 | 1.293 | 0.572 | 1.781*** | 0.327 | 2.126*** | 0.357 |
| College and above | 0.559*** | 0.037 | 0.61 | 0.344 | 2.761*** | 0.48 | 3.981*** | 0.63 |
| **MPCE Quantile: Rich®** |  |  |  |  |  |  |  |  |
| Middle | 1.15*** | 0.044 | 0.796 | 0.287 | 0.659*** | 0.076 | 0.499*** | 0.059 |
| Poor | 1.072** | 0.037 | 0.605 | 0.205 | 0.538*** | 0.059 | 0.504*** | 0.053 |
| Basic Amenities | 0.792*** | 0.057 | 2.054 | 1.637 | 1.468 | 0.36 | 1.478* | 0.337 |
| **Working Status: Never Worked®** |  |  |  |  |  |  |  |  |
| Currently Working | 1.437*** | 0.068 | 0.706 | 0.337 | 2.184*** | 0.329 | 1.381** | 0.176 |
| Currently Not Working | 1.285*** | 0.065 | 1.475 | 0.716 | 1.089 | 0.199 | 1.017 | 0.142 |
| Retired | 1.349*** | 0.105 | 1.855 | 1.276 | 3.645*** | 0.678 | 1.071 | 0.192 |
| **Self-rated health: Very Good®** |  |  |  |  |  |  |  |  |
| Good | 1.15* | 0.083 | 0.419 | 0.222 | 1.071 | 0.167 | 0.802 | 0.108 |
| Fair | 1.152* | 0.084 | 0.657 | 0.334 | 0.882 | 0.144 | 0.795* | 0.11 |
| Poor | 1.565*** | 0.125 | 0.928 | 0.546 | 1.065 | 0.209 | 0.785 | 0.138 |
| Very Poor | 1.904*** | 0.232 | 1.284 | 1.14 | 0.807 | 0.36 | 0.778 | 0.288 |
| **Morbidity: No Morbidity®** |  |  |  |  |  |  |  |  |
| Morbidity | 1.096*** | 0.038 | 1.467 | 0.465 | 1.286** | 0.128 | 1.136 | 0.109 |
| Multimorbidity | 1.001 | 0.042 | 1.098 | 0.421 | 1.36*** | 0.154 | 1.477*** | 0.15 |
| **Physical Activity Status: Never®** |  |  |  |  |  |  |  |  |
| Often | 1.004 | 0.042 | 0.948 | 0.399 | 1.062 | 0.127 | 0.818 | 0.101 |
| Frequently | 0.984 | 0.036 | 1.586 | 0.529 | 1.046 | 0.106 | 0.849* | 0.083 |
| **Natural disasters: No®** |  |  |  |  |  |  |  |  |
| Yes | 0.901 | 0.097 | 0 | 0.001 | 0.264** | 0.154 | 0.738 | 0.274 |
| **Man-made disaster: No®** |  |  |  |  |  |  |  |  |
| Yes | 1.047 | 0.14 | 1.51 | 1.538 | 1.918** | 0.563 | 1.421 | 0.457 |
| **Constant** | **0.057\*\*\*** | **0.012** | **0** | **0** | **0.005\*\*\*** | **0.003** | **0.005\*\*\*** | **0.003** |

**Note:** RRR: relative risk ratio, St. Err: standard error, ®: reference category, \*\*\* $p < .01$ , \*\* $p < .05$ , \* $p < .1$

## DISCUSSION

Health insurance is an important element for all households, as it helps people seek out their health behavior and prevents any catastrophic health expenditures. These findings, based on the nationally representative Longitudinal Aging Survey of India, shed light on the poor distribution of health insurance schemes all over the country, as a very wide number of people not seeking any health insurance came into the picture from our analyses. We focused on the near-older population aged 45 years and older, as this age group faces a greater risk of acute and chronic diseases than the rest of the population does, thus requiring benefit.

Given that we have studied the four important and very different types of health insurance, Government, Community, Employer, and Private, utilized in this country by many, the determinants considered for this study are that Government health insurance is the predominant choice of the four types of health insurance among the study participants. This could be due to different priorities and affordability concerns for the people. The insurance coverage decreases with increasing age group because of the declining sample size. Therefore, understanding age-related risk profiles is crucial for developing health insurance strategies that cater to their vulnerabilities. Males occupy a major position in providing government, employer and private health insurance than females, and females access more community health insurance than males, possibly because of their large participation in community involvement, such as self-help groups (SHGs). Hence, understanding gender-specific patterns can also be useful. The ruralCurban divide suggests potential diversity, as urban areas are more inclined toward private and employer health insurance than rural areas with more government health insurance. Notable regional differences have been shown from these two tables, where certain health insurance centers cater to places based on geographical divisions. These may reflect economic disparities as opposed to equitable coverage. The religion groups and caste categories also showed distinct preferences for health insurance. A higher educational attainment beyond a college degree, a rich household, or a household with basic amenities such as cooking fuel, drinking water, electricity, or toilets are associated with a high likelihood of having private health insurance. Currently working individuals and those who retired have a higher prevalence of employer health insurance, which strongly influences the importance of employment and its status in their choices. Individuals with very good self-rated health status, no morbidity, or who never engaged in physical activities were more likely to be without any health insurance. This defines the role of a preconceived health status and lifestyle choices in making health insurance decisions. Conversely, individuals who rated their health status as poor were inclined toward government health insurance because of affordability concerns, and individuals with multimorbidity were inclined toward private health insurance because of the feasibility of coverage.

## CONCLUSION & POLICY IMPLICATIONS

The current epidemiological transition as well as the prospective demographic transition that India will experience in the coming decades has raised serious concerns about accessibility to healthcare services. For many reasons, as mentioned in the literature, a mass population, especially a poorer population, is not able to access even primary healthcare services, which leaves a serious question of how to make these services universal and affordable. One of the prominent solutions is ‘health insurance,’ which not only makes healthcare services accessible and affordable but also helps improve healthcare facilities, as mentioned in the literature.

Our study answers the demand-side issues of various health insurances by focusing on the sociodemographic, health, and economic factors that affect the choice of four different health insurance programs among the population aged 45 and above. These factors explain the accessibility, affordability, and social factors that influence individuals’ decisions regarding being insured with a health insurance plan. Knowing these factors and reasons is very important in tracing the actual lags in the Indian health insurance market and will help policymakers formulate health insurance policies to make them attractive and useful for the mass population. In our research endeavor, we found that among the determinants we considered regarding the demographic profiles of the study population, their health situation and economic status influenced the population and described the wider fluctuations in people’s choices of health insurance, in which government health insurance tops the list followed by private health insurance.

The nuanced differences among the determinants have clear policy implications for designing and implementing appropriate health insurance programs that not only cover most of the healthcare needs of the participants but also help target the needful section of society. This approach will make the Indian health insurance market transparent and reduce the chances of failure of health insurance schemes. Future research can explore in depth the qualitative aspects of certain health insurance choices.

## Data Availability

The study utilizes a secondary source of data that is available upon reasonable request through https://www.iipsindia.ac.in/content/LASI-data

## SUPPLEMENTARY INFORMATION

## Acknowledgments

The authors gratefully acknowledge the International Institute for Population Sciences (IIPS) for providing the LASI. The study uses secondary data, which are available upon reasonable request through https://www.iipsindia.ac.in/content/lasi-wave-i.

## Authors’ contributions

Conceptualization of the paper and methodology by Madhubrota Chatterjee.

Formal analysis and supervision were provided by all the authors.

Visualization: Madhubrota Chatterjee.

Writing the original draft: Prashant Kumar Choudhary, Alok Aditya, and Madhubrota Chatterjee.

Review and editing: Prashant Kumar Choudhary, Alok Aditya, and Madhubrota Chatterjee.

## Availability of data and materials

The study utilizes a secondary source of data that is available upon reasonable request through https://www.iipsindia.ac.in/content/lasi-wave-i

## Declarations

### Ethics approval and consent to participate

The data are freely available in the public domain, and survey agencies that conducted the field survey for the data collection have collected prior consent from the respondents. The local ethics committee of the International Institute for Population Sciences (IIPS), Mumbai, confirmed that all the methods were carried out following relevant guidelines and regulations. Written informed consent was obtained from all the participants who participated in the study. All the experimental protocols were approved by the IIPS and the licensing committee. Informed consent was obtained from all the participants.

